# An exploration of healthcare providers’ learning needs and strategies for engagement in Polygenic Risk education

**DOI:** 10.1101/2025.07.17.25331741

**Authors:** Amy Clark, Courtney K. Wallingford, Jennifer Berkman, Aideen McInerney-Leo, Amy Nisselle, Bronwyn Terrill, Nathan Palpant, Mary-Anne Young, Paul James, Tatiane Yanes

## Abstract

Polygenic risk scores (PRS) provide an estimate of the genetic contribution to health conditions. Despite increasing clinical translation, healthcare providers (HPs) report a lack of PRS knowledge, representing a major barrier to safe and effective use in practice. This study aimed to i) identify HPs’ learning and resource needs for PRS delivery, and ii) outline strategies to best engage clinicians in PRS education, with findings used to inform the co-design of an educational program. To ensure informed responses, genetic healthcare providers with prior experience using PRS, and/or who had completed PRS education were recruited to participate in focus groups (n=30). Recordings were transcribed and content analysis conducted with themes mapped to the Capability, Opportunity and Motivation model for Behavior change (COM-B) to identify strategies to engage providers in PRS education. Among this cohort of experienced providers, residual PRS-related knowledge, skills and implementation gaps were frequently noted. Two themes encompassed PRS learning and resource needs: i) PRS specific knowledge base including fundamental principles, understanding clinical guidelines and test limitations, and ii) communication skills needed to discuss results and facilitate risk management and health behavior changes. Themes mapped to capability included access to training and time-poorness as a primary barrier. Limited awareness of educational initiatives, including practice resources and position statements from professional bodies, was noted. Opportunities comprised of building on existing workplace training and activities such as multidisciplinary team meetings and journal clubs. All participants noted that motivation for completing PRS training was primarily driven by a desire to improve patient-centered care and clinical outcomes. Findings highlight the complexity of PRS education and priority learning areas and will be used to inform the development of tailored PRS education for HPs to support implementation of PRS into clinical research and practice.

## Introduction

Polygenic risk scores (PRS) are an emerging genomic tool that estimates an individual’s genetic predisposition to common health conditions, based on the combined effect of many (hundreds to thousands) of genetic variants. While each variant has a small effect on health risk, their cumulative impact can stratify individuals into different levels of risk across a population. ^1^ Unlike traditional genomic testing, which typically applies to a small subset of the population, PRS can be generated for all individuals and for most complex health conditions. ^2^ PRS can be considered a patient’s intrinsic background genomic risk and can be used alongside other risk factors to inform personalized risk estimates. Polygenic risk is increasingly being integrated into clinical practice, with several commercial laboratories now delivering PRS testing. ^3,4^ Healthcare providers recognize the potential clinical utility of PRS and that testing will become integrated into future practice.^5–9^ However, they feel ill equipped to offer this test and PRS testing is not currently included in most providers training programs.^5–11^ The lack of PRS education has also been identified as a key implementation barrier, including by expert professional organisations.^12^

Genetic Healthcare Providers (GHPs, i.e., genetic counsellors and clinical geneticists) are likely to be at the forefront of PRS implementation and education of other providers. However, GHPs also lack PRS knowledge and educational opportunities, providing a barrier to education of other healthcare providers. There are limited PRS education initiatives, and GHPs’ learning needs and preferences across clinical disciplines are not well understood. ^13^ To date, PRS education has been aligned with research programs and provided only to participating healthcare providers with limited external access. While these studies increase self-reported knowledge and confidence using PRS, they were time intensive to deliver, not available beyond the study, and their clinical impact has not been evaluated.^13,14^ Thus, novel approaches to PRS education are needed to support scalable delivery methods that consider long-term changes to clinical practice.

Incorporation of stakeholder perspectives early in development of clinical education programs is essential to identify learning needs, and facilitators/barriers to engagement.^15^ Providing education alone is akin to ‘*leading a horse to water’*; additional interventions are required to deliver practice changes. The Capability, Opportunity and Motivation model of Behavior change (COM-B) offers a framework for understanding factors that enable providers to engage with, and apply new knowledge.^16^ This model posits that behavior change (i.e. engaging with PRS education) occurs when three conditions (capability, opportunity and motivation) are met. Capability includes having the necessary knowledge/skills (psychological) and physical ability (physical) to conduct the behavior. Opportunity refers to external factors that make the behavior possible (physical), and a social environment supportive of the behavior (social). Lastly, motivation describes planning and decision-making processes (reflective), and development of habits and impulses (automatic). Application of this model enables systematic identification of factors influencing uptake of education, which may translate to changes in clinical practice.

This study aimed to i) identify healthcare providers’ learning and resource needs for PRS delivery across clinical disciplines, and ii) identify strategies to best engage clinicians in PRS education. Findings will inform the development of future PRS education initiatives for healthcare providers.

## Methods

### Study design and participant recruitment

This qualitative study used focus groups to identify healthcare providers’ PRS learning needs and explore barriers/facilitators to engagement in PRS education. Eligible participants were GHPs working in Australia or the United States with PRS experience (i.e. ordering PRS tests, returning results, and/or prior PRS education). These criteria ensured informed opinions were captured from clinicians with insight into knowledge and skills required for clinical PRS use.

Various strategies were employed to facilitate recruitment. Firstly, GHPs in Australia known to have PRS experience were sent an invitation email by author AC. Study information was also distributed via professional organization newsletters and on social media in Australia and USA. A snowballing approach was employed with participants asked to share study information with colleagues. Given the initial low participation rate among clinical geneticists, targeted recruitment via study invitation to professional body mailing lists. Interested Individuals were given a link to the online consent form and a pre-study demographic survey. Interviews were held where focus groups could not be attended. While different methodologies, this approach ensured expert perspectives were captured in the study.

### Data collection

An interview guide was developed by the research team encompassing experiences of previous PRS education, learning and resources needs, and strategies to support engagement in PRS education (Supplementary Material 1). The COM-B model of behavior change^17^ informed questions around engagement with PRS education. All interviews and focus groups were conducted by authors AC and JB. Previously described methodology for focus group facilitation was applied.^18^ All focus groups and interviews were recorded, transcribed verbatim and de-identified. Participants were given a $25AUD Amazon voucher as an acknowledgement of their time. Data collection occurred February 2023 to April 2024.

### Data analysis

Focus group and interview transcripts were analyzed by author AC and content analysis conducted using NVivo 14 software.^19,20^ Firstly, three focus group transcripts were analyzed and units of meaning were assigned to participant answers. These units of meaning were then organized into learning/resource needs, or into one of the six sub-components of the COM-B model. Units of meaning and coding tree were discussed and adjusted (AC, JB, TY and CKW). All transcripts were then coded by AC with frequent meetings with JB, TY and CKW to reconcile any discrepancies in the coding process. Author JB independently re-coded 20% of the data to ensure coding accuracy, achieving a high inter-rater reliability (Kappa 80%).^21^ Illustrative quotes were selected to reflect key concepts, and findings used to identify strategies to support PRS education and subsequent use in clinical practice.

## Results

### Demographics

Of the 65 GHPs who expressed interest in the study, 12 were ineligible due to no prior PRS experience. Of the 53 eligible individuals, 30 participated (n=21 via focus groups, and n=9 interviews). Reasons for non-participation included: no response to subsequent invitation (n=9), incorrect contact details provided (n=11), and inability to co-ordinate a mutually convenient time (n=3). Focus groups were 65-77 minutes and interviews 32-48 minutes. Most participants were females (n=25, 83%), genetic counsellors (n=25, 83%), from Australia (n=26, 87%), and with prior PRS experience in the breast cancer setting (n=14, 47%) (Table 1). Mean age was 40 years (range 24-67 years), with similar distributions across years of practice and time working with PRS. Most participants reported attending previous education and or gaining experience with PRS through recruitment for clinical research (n=21, 70%).^13,22,23^

**Table 1:** Participant demographic characteristics and prior experience using PRS.

**Table 2:** Summary of genetic healthcare providers’ learning and resources for PRS.

| Theme | Quote |
| --- | --- |
| **PRS specific knowledge base** |  |
| Not standard training for GHPs | *"I trained over a decade ago, and we unfortunately had very little true formal GWAS training...there's very little of that training or education for genetic counsellors. Because we generally haven't had to use or decide to use screening tests...We've ordered diagnostic gene tests and let the clinicians handle the mammograms and the cholesterol panels..."* (Participant 24, Genetic Counsellor) |
| Fundamental knowledge: GWAS, PRS development, evaluation and integrated risk | *"I like understanding the first principles or the kind of underlying basis of whatever's sort of being calculated. Like not the, not an enormous detail, but just enough so that you kind of have a rough idea about how these things are calculated."* (Participant 2, Clinical Geneticist)<br>*"I think there needs to be some foundational education about things like model discrimination, internal and external validation"* (Participant 24, Genetic Counsellor) |
| PRS considerations | *"I guess the question that the problem needs to be asked with an educational point of view, well what's it being used for?"* (Participant 27, Clinical Geneticist)<br>*"...how we use the polygenic risk in terms of the different ethnicities and how we deal with that and how I discuss that with the patient".* (Participant 1, Genetic Counsellor)<br>*"...having an understanding of whether or not that [missing SNPs] will actually influence the accuracy of the risk assessment, I think is really important...I'd sort of assume to read in these reports..."* (Participant 17, Genetic Counsellor) |
| Emerging evidence | *"I have the impression...we are kind of at that we have clinical validity, but not necessarily [clinical] utility and that's kind of what we're figuring out, isn't it?"* (Participant 2, Genetic Counsellor) |
| Clinical guidelines | *"Without guidelines, it's hard to convince people that this is something they need to do."* (Participant 27, Clinical Geneticist)<br>*"Do I put this woman on insulin because she's freaking out about her slight increased risk of diabetes? And that's a really good question. What needs to be done? What criteria, what guidelines?"* (Participant 20, Genetic Counsellor) |
| Level of knowledge required | *"I sit somewhere in between wanting to know absolutely everything and absolutely nothing about a PRS. There are some technical details that I need to know when I'm giving a result. If something doesn't make sense to me on a report, I will go to the lab and say, you need to explain this because it doesn't make sense why there looks like they've got a really high PRS, you've reported them as moderate...and they help explain that. But do I ever really want to do my own PRS score? Probably not."* (Participant 28, Genetic Counsellor)<br>*"If I can understand how something is generated, then it helps me to be able to communicate it."* (Participant 8, Genetic Counsellor)<br>*"There will be a kind of subgroup of patients who are really quite interested in how a PRS is calculated and having* |
|  | *that knowledge up my sleeve will be useful.* (Participant 16, Genetic Counsellor) |
| Direct clinical implications | *"What I really gained from doing the PRiMo [PRS] training...was a really kind of practical breakdown of, well, if this is, this is the information we're gonna provide you on a report and then being able to translate that into useful clinical relevant information that the patients can then use to inform their risk management, or risk perception."* (Participant 13, Genetic Counsellor) |
| **Communication skills** |  |
| GHP are already trained in the skills needed for PRS communication | *"You need to be able to sense the level of your client's understanding. You need to be able to work out what language to use with them. You need to be able to use those active listening skills and reflective statements to try and figure out, are they with me? They're all the same skills that you use, no matter what you're counselling about."* (Participant 19, Genetic Counsellor) |
| Complex inheritance discussion | *'It can become quite a complex discussion and that is quite difficult when someone has inherited the familial mutation. So, you've given them a result that has huge emotional implications and then trying to discuss something quite complex with them.'* (Participant 3, Genetic Counsellor) |
| Practice-based examples and resources to support PRS communication | *"We all pick up useful analogies, or ways of phrasing things as we go through our training, and if we can concentrate that into a [PRS] training course, so that we don't need to observe other people doing it, that'd be amazing."* (Participant 2, Clinical Geneticist)<br>*"...experiential learning is my better preferred model and if I've had the chance to role play something a bit new as opposed to watching a role play, I'm probably more likely to be able to try it out ... until I actually have to do it, that's not putting me on the spot and seeing if I'm fumbling over my words or I really don't understand this."* (Participant 26, Genetic Counsellor) |
| Potential for behaviour change | *"I'm really interested in the combined [score] because it seems to be a little bit more powerful. It also has the potential for people to see where the risk factors are and how to change them and it gives them back power around some of these conditions."* (Participant 28, Genetic Counsellor) |
| GHP do not feel equipped to influence behaviour change | *"It's [behaviour change] is not something I ever had experience in communicating. It's not something that was touched on in my training at all. So, it was entirely new language phrasing."* (Participant 14, Genetic Counsellor) |
|  | *"... my ability to change someone's entire behaviour with a one-off 20-minute conversation is very minimal."* (Participant 14, Genetic Counsellor) |

### Learning needs

Frequently, participants recognized a training gap regarding PRS and screening tests. They acknowledge that current clinical training focuses on diagnostic genomic testing, which differs from the epidemiological concepts used in PRS and population risk stratification. Overall, there was a consensus that healthcare professionals should understand the concept of genome-wide association studies (GWAS), how GWAS informs PRS development, integrated risk, and epidemiological methods of evaluating risk assessment models. Additional PRS considerations included understanding clinical context for test use, impact of ancestry on result interpretation, and confidence in the score. Regarding clinical use, participants frequently recognized the strong evidence for clinical validity, modelling data supporting PRS, and potential clinical applications. They were aware that clinical trials are still building evidence for real-world clinical utility, limiting the development of PRS and integrated risk-specific clinical guidelines. The lack of clinical guidelines was viewed as a primary barrier for implementation, prompting participants to note that future PRS education may have limited value without guidelines to inform patient risk management.

Views varied about the depth of knowledge required by providers in statistical and epidemiological processes related to PRS. Most participants felt a simplified gist understanding of these processes was sufficient to interpret reports, identify potential issues, communicate with laboratories and deliver results. Conversely, a subset of participants wanted extensive background information on PRS development, statistical modelling and evaluation. For these individuals, having detailed knowledge of the technical aspects of PRS enhanced their capacity to interpret and communicate information to patients. Regardless of the level of background knowledge, participants agreed that PRS education should have a strong focus on direct clinical implications for patients and a practical breakdown of tasks such as report/result interpretation and risk communication.

Nearly all participants felt that GHPs already had the necessary skills to communicate complex risk information. These skills included capacity to distil complex information for patients, active listening, reflective practice and exploring the psychological and familial implications of results. However, participants emphasized the need for education and resources to help communicate integrated risk, i.e. combining polygenic, monogenic and non-genetic risk factors. They reported challenges explaining both Mendelian and polygenic inheritance, as well as providing multiple risk periods (i.e. 5-year, 10-year and lifetime risks). Participants suggested practical tools such as example explanations, analogies, common patient questions, and video role plays to support communication. Several participants also noted the power of integrated risk scores in motivating healthier patient behaviors, but they felt undertrained in strategies to affect change, particularly within the context of single results appointments.

### Factors influencing engagement with PRS education

This study used the COM-B model of behavior change to comprehensively map the factors impacting GHP engagement with PRS education. Results are summarized in Table 3.

**Table 3:** Capabilities, Opportunities and Motivations that facilitate clinicians building their PRS knowledge and skills.

| Theme description | Representative quote | Implication for training |
| --- | --- | --- |
| **Capability: psychological (knowledge and mental capacity)** |  |  |
| Building upon existing knowledge base | *"It's one of the real challenges...of most workshops that I've been to where people just haven't been able to accommodate that diversity of expertise."* (Participant 2, Clinical Geneticist) | Customizable education to accommodate varied baseline knowledge levels and learning needs. |
| Awareness of existing education | *"I don't know if that resource has been integrated into our training curriculum yet. I think it's listed...as a supplemental resource that students are encouraged to check out...But honestly, it sounds bad, but I feel like nobody else knows about the [NSGC] practice resource"* (Participant 24, Genetic Counsellor) | Dissemination and visibility of resources to improve knowledge of training resources<br>Development of central PRS resource hub. |
| **Capability: physical (physical capacity)** |  |  |
| Accessibility - ability to attend or access existing education programs/initiatives. | *"it's really hard for me to attend an actual place to do training...[anything] that makes things way more accessible for me whenever via, you know, telehealth or virtual or something I can do online in my own time."* (Participant 18, Genetic Counsellor) | Ensuring accessible education |
| Time poorness - finding time to complete education. | *"...having smaller modules is really good because, you know, we're really busy. Sometimes you've got a spare half hour or a spare hour and being able to just break it down and do a small little component one day and then maybe another hour another day makes it a little bit more feasible."* (Participant 8, Genetic Counsellor) | Micro learning, chunking information. |
| **Opportunities: Physical (inanimate/physical environment)** |  |  |
| Integration of PRS education in current workplace education | *"We've ended up talking about it in a couple of our supervision sessions. That's actually been really helpful to really just like, throw it out there. These are the experiences, these are the challenges we have, and getting that group feedback and ideas, has been really supportive."* (Participant 14, Genetic Counsellor) | Embedding PRS education into MDTs, journal clubs, supervision and grand rounds. |
| Education initiatives that facilitate expert and peer case discussion | *"...the national variant review meeting...people bring their cases. All the labs are there, all the services are there. You work through these tough variants, and you come away with something and it's like actually super worthwhile."* (Participant 3, Genetic Counsellor) | Online case discussion meetings with subject matter experts. |
| **Opportunities: Social (social environment/culture)** |  |  |
| Workplace culture that values PRS education | *'Our unit did it as a group. So that made it very easy... it's actually something that's recommended by your workplace... you're doing it with* | Demonstration of importance through peer promotion (e.g. inclusion of |
|  | *people, you know and feel comfortable asking questions.'* (Participant 1, Genetic Counsellor) | PRS-focused sessions in conferences). |
| Institutional support for PRS education | *'I think that's where we struggle the most. It always ends up being something that we tack on or squeeze in...unless we have a, a way of actually future proofing it or planning it.'* (Participant 8, Genetic Counsellor) | Identifying how PRS training aligns to health system policies and strategic plans.<br>Blocked time, funded education, workload considerations, upskilling focus aligned with strategic priorities. |
| **Motivation: Automatic (Habits and impulses)** |  |  |
| Desire to provide personalised care for patients | *"...once you look at [lifestyle] factors and someone realises that they have control over some of those factors, it's a game changer... They feel like there's real, power that's been given to them to know this information and for some reason it is way less scary than doing germline... PRS feels like it's much more in control."* (Participant 28, Genetic Counsellor) | Highlight individual clinical and personal utility of PRS in educational program. |
| Consistently encountering PRS results or referrals drives a need to upskill | *"At the beginning it kind of fell in my lap... I ended up seeing a bunch of referrals for that reason. So, I started to educate myself, just trying to figure out what to tell the patients in that space."* (Participant 23, Genetic Counsellor) | Identify examples of referral/case scenarios in educational resources (i.e. case-based learning)<br>Consider patient needs e.g. frequently asked questions by patients |
| **Motivation: Reflexive (conscious thought and planning)** |  |  |
| Facilitating consistent care across services | *"... I don't know what we would do with if this becomes mainstream clinical... what we do with families with moderate risk gene... And then when we start getting families with different risk information for different family members... from an industry perspective, some sort of agreement about how we're going to deal with that when it comes."* (Participant 7, Genetic Counsellor) | Develop PRS model of care alongside educational programs to ensure consistent evidence-based practice. |
| Maintaining professional standards | *'It was the feeling that PGS was going to be inevitable, and I better learn about it now or I'll be left behind.'* (Participant 3, Genetic Counsellor)<br>*"I think to get them involved is to say, this counts towards your CPD... And it's kind of killing two birds with one stone that they're kind of getting a trade-off for learning and being educated in this space. And then maybe that hooks people in and they say, 'oh, actually I really like this and this is something that I want to keep learning about'."* (Participant 25, Genetic Counsellor) | Recognising PRS education contributes to CPD and professional standing |

CPD = continuing professional development

#### Capability

Two overarching concepts were identified relevant to psychological capability: i) customized learning depending on the individual’s knowledge base and ii) dissemination and awareness of educational resources. When reflecting on prior professional development, participants noted limited consideration of varied knowledge levels and little accommodation of different learning styles, which negatively impacted subsequent engagement. Lack of awareness of existing educational resources was noted as a barrier to engagement, with some participants unfamiliar with practice resources supplied by professional bodies. Participants suggested that collating PRS resources into a central PRS learning hub may facilitate access and improve dissemination of resources.

Participants also identified physical capability barriers including i) accessibility issues and ii) being time poor. Several respondents described needing to request permission to access restricted educational materials used in research studies. Participants suggested time constraints could be addressed with flexible modes of delivery such as chunking content into micro-lessons. However, they also recognized the potential limitations of web-based programs, including non-completion and disengagement due to information density and distraction.

#### Opportunity

Participants described potential enhancements to physical opportunities that included i) building PRS education into existing workplace training and ii) facilitating case discussion with peers and experts. Adapting current continuing professional development (CPD) activities to incorporate PRS, rather than additional programs, was highly valued by participants. Clinical supervision sessions, multidisciplinary team (MDT) meetings and journal clubs were identified as settings to incorporate PRS education. Additionally, participants strongly valued access to experts within MDT’s, journal clubs, and research study teams which facilitated peer-to-peer discussion of challenging cases, and the development of PRS-related skills. Lastly, based on the value of peer-to-peer learning, participants suggested developing forums for PRS interpretation where complex PRS cases could be submitted, with expert feedback provided, facilitating reciprocal learning.

Factors impacting social opportunities to engage in PRS included: i) workplace culture/values toward PRS education, and ii) institutional support. Participants emphasized that supportive workplace cultures valuing professional development and collective learning fostered engagement in education about emerging technologies. Those involved in PRS implementation studies noted that shared commitment to evidence-based practice, inclusive team training and prioritizing time for learning were key factors enabling their participation in training. Furthermore, participants indicated a lack of institutional support (e.g. protected time and funding for staff training) as a major barrier to engagement in PRS education. One suggestion to secure institutional support was to align the learning outcomes of local education initiatives with organizational strategic priorities. While this idea was not frequently raised, it generated much interest and discussion during focus groups.

#### Motivation

Automatic motivation to engage in PRS education was primarily driven by a desire to improve patient-centered care. Specifically, nearly all participants felt that PRS could enhance personalized care, provide answers for families about unexplained heritability (i.e. when no causal pathogenic variant is identified despite a significant family history), and/or provide more accurate risk assessments. Motivation to engage in PRS use was further driven by patient referrals related to PRS. Thus, GHPs wanted to engage with PRS education to build their confidence and capability in applying PRS in clinical practice.

Factors that influenced participants’ reflective motivation included: i) upskilling in PRS as a professional standard and ii) wanting to facilitate consistent care for families. GHPs wanted to maintain professional standards and stated that they “*do not want to be left behind*” in what was described as the next frontier in genomic medicine. In some cases, participants felt that having skills with PRS communication would improve their employability, and that attaching CPD points to educational activities would be beneficial. Lastly, participants wanted to ensure consistent PRS case management and suggested this could be best facilitated through practice-based group training.

## Discussion

This study explored GHPs’ learning needs and factors influencing their engagement in PRS education. Preference for patient-centered PRS education was a key finding, with providers emphasizing risk communication and clinical management over comprehensive knowledge of PRS development and evaluation. Overall, GHPs favored customizable, case-based learning, and resources that included example explanations and responses to common patient questions. Based on study findings, specific suggestions for future PRS education are proposed, particularly resource differentiation to accommodate varied learning needs, flexible modules and accessibility through development of a central PRS resource hub (Table 3). Time constraints, competing workloads and lack of systemic support were major barriers to completing PRS education. Integrating PRS education into workplace training, such as journal clubs, supervision and case discussions were suggested avenues to address these barriers. PRS represents a new landscape in healthcare provider education and practice. Understanding PRS development requires analytical and statistical skills, such as interpreting risk estimates, evaluating model performance and understanding the implications of population genetics.^24^ All participants felt that a foundational knowledge in these areas was essential, though the level of engagement may vary. Currently, these skills are not explicitly included in clinical practice frameworks or training competencies outlined by professional bodies such as the Human Genetics Society of Australasia (HGSA) or the National Society of Genetic Counsellors (NSGC).^10,11^ As a result, many clinicians may lack formal training in these areas, highlighting a critical gap in preparing the workforce for the integration of PRS into clinical care.

While GHPs were highly motivated to learn about PRS to personalize patient care, the lack of PRS-specific clinical guidelines has been described as demotivating for broader groups of clinicians.^7^ Clinical guidelines are seen as necessary to support clinicians in the advice they provide to patients. Providers recognized that there has not been sufficient time to generate long-term evidence for clinical utility, which could be used to develop clinical guidelines.^25^ This evidence gap presents a significant challenge for PRS education, as providers have limited access to clear information about clinical and lifestyle/behavioral interventions, which can be offered to patients receiving PRS results. In the absence of PRS-specific guidelines, clinicians need to consider the specific setting in which PRS is being applied and determine whether existing guidelines are applicable. For example, comprehensive risk management guidelines based on absolute risk thresholds exist for some common cancers.^26–28^ These can be directly applied to PRS-based risk estimates. However, in other settings such as psychiatric conditions, or PRS for newborn screening for type one diabetes, new guidelines will need to be developed.^29,30^

Participants noted that their current skillset in communicating complex risk information was transferable to the PRS setting. However, skill gaps were identified in the communication of integrated risk information. Participants also identified challenges in delivering different risk time frames, such as 5-year, 10-year and lifetime risk figures. Given preferences for patient-centered education, future work should prioritize the development of point-of-care resources, such as visual aids and patient-friendly reports to facilitate risk communication. While visual aids, such as ‘the jar model’,^31^ have been adapted for polygenic testing, assessment of their utility has been limited beyond psychiatric genetics.^32^ Future research should aim to explore how other existing resources can be adapted to PRS settings such as familial cancer and cardiac genetics. Furthermore, educational resources could include examples of metaphors and analogies, tools which are known to aid communication of complex genetic risk information.^33^

Identification of facilitators and barriers to engagement in clinical PRS education allows for the development of wholistic clinical education strategies. To optimize engagement in PRS education initiatives, GHPs expressed a need for customizable and flexible education, with a preference for practice-based learning, consistent with other genomics education studies.^34–36^ Practice based learning aligns with adult learning theory, which highlights the involvement of the participant in planning and evaluation of their learning, as participant experience, immediacy, and problem-based learning are integral to engagement.^37^ These educational strategies are associated with improved physician performance and patient outcomes.^38,39^ Furthermore, participants noted limited time, competing workloads, lack of institutional support, and lack of awareness of existing education resources as barriers for PRS education engagement, challenges well documented internationally.^40,41^ This highlights the need for implementation strategies that promote institutional support^40,41^ and broaden reach through centrally located education initiatives (e.g. a designated PRS education website).^14^ Future research should consider evaluation approaches during development of new PRS education initiatives, as few genomic education interventions are evaluated beyond immediate measures of knowledge and attitudes.^42^

Limitations of this study included the predominance of female genetic counsellors from Australia, with experience in the cancer setting. While this sample is representative of the Australasian GHPs profile,^43^ views regarding use of PRS among other clinicians in Australia and internationally may vary based on specialty, area of practice and other demographical factors. Conversely, recruitment of GHPs with experience in PRS enhanced discussions and explorations of learning needs that were informed by clinical practice. Future studies should aim to explore views of naïve users, underrepresented GHP groups and potential users in other specialties. Strengths of this study include the use of a theoretical framework (COM-B model) to comprehensively identify the barriers and facilitators impacting engagement in PRS education.

This study identified GHPs’ PRS education learning needs focusing on the fundamentals of PRS development, integrated risk communication and clinical implications. To enhance clinician engagement, PRS education and training modules should be customizable, flexible, and accessible, integrated into existing work-based training, and delivered in team settings with employer support. In alignment with providers’ desires to facilitate personalized care for patients, clinical guidelines and standardized care models should be developed to inform management based on PRS and integrated risk assessments.

## Data Availability Statement

The de-identified data supporting study findings are available for the purpose of verifying or contextualizing our conclusions. Requests should be sent to the corresponding author describing the requester’s qualifications and the reason for the request.

## Supporting information

Interview guide

## Acknowledgements

This study was conducted by Amy Clark (student investigator) in partial fulfilment of the requirements of the Master of Genetic Counselling degree from the University of Technology, Sydney. The authors would like to extend thanks to the participants in this study.

## Author Contribution

Conceptualization: T.Y. and A.C. Data curation: A.C., J.B. and T.Y. Formal analysis: A.C., J.B. C.K.W. and T.Y. Funding acquisition: T.Y., P.J and M.Y.A. Investigation: A.C., J.B. and T.Y. Methodology: T.Y., A.M.L., L.D., A.C., J.B., A.M., B.T. and C.K.W. Project administration: T.Y., A.C., and J.B. Supervision: A.M.L, L.D and T.Y. Validation: J.B. and T.Y. Visualization: T.Y., A.C., J.B. and C.K.W. Writing original draft: T.Y., A.C., J.B. and C.K.W. Writing – review and editing: J.B., C.K.W., A.N., B.T., MA.Y., L.D., A.M.L., P.J. and T.Y.

## Ethics Approval

The study was approved by the University of Queensland Human Research Ethics Committee (2022/HE001551) and the University of Technology Sydney Human Research Ethics Committee for ratification (ETH22-7717). All participants provided informed consent for the study.

## List of tables, figures, and Supplementary material

Supplementary Material 1: Appendix E Interview guide

## References

1. Xiang R, Kelemen M, Xu Y, et al. Recent advances in polygenic scores: translation, equitability, methods and FAIR tools. Genome medicine. 2024;16(1):33–14.

2. Yanes T, McInerney-Leo AM, Law MH, Cummings S. The emerging field of polygenic risk scores and perspective for use in clinical care. Human Molecular Genetics. 2020;29(R2):R165–R176.

3. Natarajan P. The first risk factor: Polygenic risk scoring for coronary heart disease. Journal of the American College of Cardiology. 2018;72(16):1894–1897.

4. Lewis ACF, Green RC. Polygenic risk scores in the clinic: new perspectives needed on familiar ethical issues. Genome Medicine. 2021;13(1):14.

5. Reddi HV, Wand H, Funke B, et al. Laboratory perspectives in the development of polygenic risk scores for disease: A points to consider statement of the American College of Medical Genetics and Genomics (ACMG). Genet Med. 2023;25(5):100804–100804.

6. Young M-A, Yanes T, Cust AE, et al. Human Genetics Society of Australasia Position Statement: Use of Polygenic Scores in Clinical Practice and Population Health. Twin Res Hum Genet. 2023;26(1):40–48.

7. Smit AK, Sharman AR, Espinoza D, et al. Knowledge, views and expectations for cancer polygenic risk testing in clinical practice: A cross-sectional survey of health professionals. Clinical Genetics. 2021;100(4):430–439.

8. Connolly JJ, Berner ES, Smith M, et al. Education and electronic medical records and genomics network, challenges, and lessons learned from a large-scale clinical trial using polygenic risk scores. Genetics in Medicine. 2023;25(9):100906.

9. Lewis ACF, Perez EF, Prince AER, et al. Patient and provider perspectives on polygenic risk scores: implications for clinical reporting and utilization. Genome Med. 2022;14(1):114.

10. Australasia HGSo. HGSA Competency Standards for Genetic Counsellors. Human Genetics Society of Australasia. https://hgsa.org.au/common/Uploaded%20files/pdfs/policies%2C%20position%20statements%20and%20guidelines/genetic%20counselling/Competency%20Standards%20for%20GCs.pdf?utm_source=chatgpt.com. Published 2022. Accessed 28 May 2025.

11. Counselors NSoG. Institutional-Based Model Credentialing Policy for Genetic Counselors. National Society of Genetic Counselors. https://www.nsgc.org/portals/0/docs/about/NSGC%20Model%20Institutional%20Credentialing%20Policy%20and%20Procedure.pdf. Published 2017. Accessed 28 May 2025.

12. Purvis R, Forrest LE, Young M-A, Limb S, James P, Taylor N. Defining next steps in the clinical implementation of polygenic scores: A landscape analysis of professional groups’ perspectives. Genetics in Medicine. 2025:101414.

13. Yanes T, Wallingford CK, Young M-A, et al. Development and evaluation of a novel educational program for providers on the use of polygenic risk scores. Genet Med. 2023;25(8):100876–100876.

14. Rasmussen LV, Overby CL, Connolly J, et al. Practical considerations for implementing genomic information resources Experiences from eMERGE and CSER. Appl Clin Inform. 2016;7(3):870–882.

15. Nisselle A, Janinski M, Martyn M, et al. Ensuring best practice in genomics education and evaluation: reporting item standards for education and its evaluation in genomics (RISE2 Genomics). Genet Med. 2021;23(7):1356–1365.

16. Michie S, van Stralen MM, West R. The behavior change wheel: a new method for characterizing and designing behaviour change interventions. Implement Sci. 2011;6:42.

17. Michie S, van Stralen MM, West R. The behavior change wheel: A new method for characterizing and designing behavior change interventions. Implementation science : IS; Implement Sci. 2011;6(1):42.

18. Krueger RA, Casey MA. Focus groups : a practical guide for applied research. 5 ed. Thousand Oaks, California: SAGE; 2015.

19. Hsieh H-F, Shannon SE. Three Approaches to Qualitative Content Analysis. Qual Health Res. 2005;15(9):1277–1288.

20. Ralph IG. Qualitative Data Analysis Software: NVivo. Qualitative research journal. 2004;4(2):77–81.

21. McHugh ML. Interrater reliability: The kappa statistic. Biochem Med (Zagreb). 2012;22(3):276–282.

22. Lennon NJ, Kottyan LC, Kachulis C, et al. Selection, optimization and validation of ten chronic disease polygenic risk scores for clinical implementation in diverse US populations. Nat Med. 2024;30(2):480–487.

23. McInerny S, Mascarenhas L, Yanes T, et al. Using polygenic risk modification to improve breast cancer prevention: study protocol for the PRiMo multicenter randomized controlled trial. BMJ Open. 2024;14(8):e087874.

24. Babb de Villiers C, Kroese M, Moorthie S. Understanding polygenic models, their development and the potential application of polygenic scores in healthcare. J Med Genet. 2020;57(11):725–732.

25. Kumuthini J, Zick B, Balasopoulou A, et al. The clinical utility of polygenic risk scores in genomic medicine practices: a systematic review. Hum Genet. 2022;141(11):1697–1704.

26. NSW CIo. eviQ. https://www.eviq.org.au/. Published 2024. Accessed.

27. Excellence NIfHaC. Published: Guidance, quality standards and advice.. https://www.nice.org.uk/guidance/published. Published 2024. Accessed.

28. Australia C. National Cancer Control Indicators. Screening. https://ncci.canceraustralia.gov.au/screening. Published 2024. Accessed.

29. Austin JC. Re-conceptualizing Risk in Genetic Counseling: Implications for Clinical Practice. J Genet Couns. 2010;19(3):228–234.

30. Kredo T, Bernhardsson S, Machingaidze S, et al. Guide to clinical practice guidelines: The current state of play. International journal for quality in health care. 2016;28(1):122–128.

31. Austin JC. Evidence-based genetic counseling for psychiatric disorders: A road map. Cold Spring Harb Perspect Med. 2020;10(6):a036608.

32. Cook CB, Slomp C, Austin J. Parents’ perspectives, experiences, and need for support when communicating with their children about the psychiatric manifestations of 22q11.2 deletion syndrome (22q11DS). J Community Genet. 2022;13(1):91–101.

33. Sexton A, James PA. Metaphors and why these are important in all aspects of genetic counseling. J Genet Couns. 2022;31(1):34–40.

34. Morrow A, Chan P, Tiernan G, et al. Building capacity from within: qualitative evaluation of a training program aimed at upskilling healthcare workers in delivering an evidence-based implementation approach. Translational Behavioral Medicine. 2022;12(1).

35. Crellin E, McClaren B, Nisselle A, Best S, Gaff C, Metcalfe S. Preparing Medical Specialists to Practice Genomic Medicine: Education an Essential Part of a Broader Strategy. Frontiers in genetics. 2019;10:789–789.

36. McClaren BJ, Crellin E, Janinski M, et al. Preparing Medical Specialists for Genomic Medicine: Continuing Education Should Include Opportunities for Experiential Learning. Front Genet. 2020;11:151–151.

37. Knowles MSHEFSRA. The adult learner: the definitive classic in adult education and human resource development. 6th Edition ed: Butterworth-Heinemann; 2011.

38. Cervero RM, Gaines JK. The Impact of CME on Physician Performance and Patient Health Outcomes: An Updated Synthesis of Systematic Reviews. J Contin Educ Health Prof. 2015;35(2):131–138.

39. Kolb DA. Experiential learning : experience as the source of learning and development. Second edition. ed: Pearson Education; 2014.

40. Pettersson F, Olofsson AD. Implementing distance teaching at a large scale in medical education: A struggle between dominant and non-dominant teaching activities. Education and information technologies. 2015;20(2):359–380.

41. O’Doherty D, Dromey M, Lougheed J, Hannigan A, Last J, McGrath D. Barriers and solutions to online learning in medical education - An integrative review. BMC Med Educ. 2018;18(1):130–130.

42. Nisselle A, Janinski M, Martyn M, et al. Ensuring best practice in genomics education and evaluation: reporting item standards for education and its evaluation in genomics (RISE2 Genomics). Genetics in Medicine: Official Journal of the American College of Medical Genetics. 2021;23(7):1356–1365.

43. Kanga-Parabia A, Mitchell L, Smyth R, et al. Genetic counseling workforce diversity, inclusion, and capacity in Australia and New Zealand. Genet Med Open. 2024;2(Suppl 2):101848.

