## Supplementary material for "An exploration of healthcare providers’ learning needs and strategies for engagement in Polygenic Risk education": Interview guide

***Interview Guide: Exploration of learning needs and preferences of genetics health care professionals in Polygenic Score education.***

**Introduction Script**

- Thank participant for agreeing to take part in this study.
- Briefly introduce the researcher and brief professional background.
- Introduce the study and briefly discuss its purpose.
- Discuss how focus group facilitator/interviewer role: to raise topics for discussion and then to listen as participants share views and experiences. Group facilitator may also encourage discussion between participants in focus groups to explore similarities and differences. Facilitator may also use brainstorming tools such as the Zoom whiteboard to prompt discussion.
- Set zoom protocols: may stay off mute and jump in or raise hand. In some discussions you will be asked to pass to another participant. Please be mindful of allowing everyone to have a say. We encourage you to build on other comments.
- Reassure participants that they are free to talk about any aspect of their experience or attitudes. There are no right or wrong or even typical answers to any of the questions that we will discuss. All views will be treated with equal relevance to the research.
- Remind participants that, with their permission, the session/interview will be recorded.
- Reassure confidentiality and the participant’s right to leave the focus group or stop the interview at any time.
- Clarify that the focus group will take approximately 1-1.5 hours and interview will take approximately 30 minutes.
- Ask whether participants have questions before commencing. **START RECORDING.**

**Focus group date:**

**Number of attendees:**

**Notes:**

**Questions**

1. Can you introduce yourself and tell me about your experience with communicating PGS?

*Prompts e.g.:*

- *Used PGS in research or clinic?*
- *How much experience do you have?*

2. Can you describe your experience receiving education in PGS? If you have not attended training, how have you gone about upskilling yourself for working with PGS?

*Prompts:*

- *What was covered in the program?*
- *What worked well?*
- *What did not work well?*

3. What are the essential skills you think genetics health care professionals need to work effectively with PGS?

*Prompt:*

- *By the end of this training, I will be able to....*
- *What don’t HCP’s need to know? Where is the line?*

4. What resources do you believe would be beneficial in supporting your education of PGS?

*Prompts e.g.:*

- *Fact sheets, flow charts, Standard Operation Procedures*
- *Are there any resources you currently use that could be adapted for PGS education?*

5. What motivated you to engage with PGS education? If you have not completed education, what would motivate you to?

*Prompt:*

- *E.g. peer support/institutional support, patient need, being prepared for emerging technology?*

6. What opportunities help you to engage in PGS education? If you have not completed education, what opportunities would help you?

*Prompt:*

- *E.g. access to funding/resources to complete training, peer/institutional support,*	*incentives?*

7. How would you prefer to engage in ongoing professional educational activities more broadly (i.e., not just PGS)?

*Prompts:*

- *Point of care - in person from experts or peers, as part of clinical practice, experiential, as part of supervision,*
- *In person education - seminars, webinars, short courses*
- *Online/Self-directed learning – e.g. YouTube – Professional organisations – google it?*
- *Do you ever learn from Social media e.g. Twitter, TikTok etc.*

8. Can you reflect on a successful education activity or activities and describe some key features?

*Prompt:*

- *How did they help you change your practice?*

9. Is there anything further you would like to add regarding PGS education?

**Interview debriefing and closure script**

- Thank the participant for sharing their experiences.
- Reshow slide 1 with contact details – if they wanted to talk with research team about anything further.
- Confirm they will receive a $25 Amazon voucher via email.
